# A quantitative framework to define the end of an outbreak: application to Ebola Virus Disease

**DOI:** 10.1101/2020.02.17.20024042

**Authors:** Bimandra Adiputra Djaafara, Natsuko Imai, Esther Hamblion, Benido Impouma, Christl Ann Donnelly, Anne Cori

**Author notes:** **Correspondence** MRC Centre for Global Infectious Disease Analysis, School of Public Health, Imperial College London, Medical School Building, Norfolk Place, London, W2 1PG.

## Abstract

Declaring the end of an outbreak is an important step in controlling infectious disease outbreaks. An objective estimation of the probability of cases arising in the future is important to reduce the risk of post-declaration flare-ups. We developed a simulation-based model to quantify that probability. We tested it on simulated Ebola Virus Disease (EVD) data and found this probability was most sensitive to the instantaneous reproduction number, the reporting rate, and the delay between symptom onset and recovery or death of the last detected case. For EVD, our results suggest that the current WHO criterion of 42 days since the outcome of the last detected case is too short and very sensitive to underreporting. The 90 days of enhanced surveillance period after the end-of-outbreak declaration is therefore crucial to capture potential flare-ups of cases. Hence, we suggest a shift to a preliminary end-of-outbreak declaration after 63 days from the symptom onset day of the last detected case. This should be followed by a 90-day enhanced surveillance, after which the official end-of-outbreak can be declared. This corresponds to less than 5% probability of flare ups in most of the scenarios examined. Our quantitative framework could be adapted to define end-of-outbreak criteria for other infectious diseases.

## Introduction

Declaring the end of an outbreak (EO) is a critical programmatic step in outbreak response. Any infectious disease outbreak can be devastating for affected populations and areas as alongside burden from infections and mortality, the outbreak status of a region or a country can influence other vital sectors such as: social, economic, political, and security (1–3). In defining the EO, it is crucial to consider the risk of cases arising in the future using objective quantitative methods. A well-timed declaration of EO is essential as it would allow affected countries to address problems in other sectors, reallocate healthcare resources to cover other important public health issues, and start post-outbreak recovery efforts.

Various definitions or criteria of an EO have been used in different outbreaks depending on the type of disease and the institutions providing technical guidance or in charge of controlling the outbreak. We retrieved examples of EO criteria from various pathogens and institutions from the literature (**Table 1**). Most of these criteria are obtained from the World Health Organization (WHO) guidelines or decided by a government’s Ministry of Health or public health authorities. The most commonly used EO criterion is a period of twice the longest incubation period without observing any new cases since the last possible transmission event (4–12). In an Ebola Virus Disease (EVD) outbreak context, the EO of EVD is declared after 42 days (twice the longest incubation period) has passed since the outcome of the last case. There are several possible scenarios of the outcome of the last case (8). If the last case is a laboratory-confirmed case, the outcome can be the second PCR negative test of blood samples or a safe burial if the person died. Additionally, if the last case is a contact of a confirmed case, the outcome is a safe burial after the person’s death.

**Table 1.** End-of-an-outbreak criteria from past infectious disease outbreaks

| **End-of-outbreak criteria definition** | **Outbreak(s) which used this criteria (References)** |
| --- | --- |
| Twice the longest incubation period of no cases since the death/recovery of the last confirmed case | Marburg (Uganda 2017) (4); MERS (South Korea 2015 & 2018) (5,6); Diphtheria (Indonesia 2017-2018) (7); Ebola virus disease (West Africa 2013-2018) (8–12) |
| Twice the longest incubation period of no cases since the reporting day of the last confirmed case | Lassa (Benin 2016) (22) |
| No new cases reported for six months | Yellow fever (DRC and Angola 2015-2017) (23) |
| No new cases reported for seven weeks | Cholera (South Sudan 2017-2018) (24) |
| No cases due to the outbreak strain reported for three months and incidence rate in the past two months dropped to the pre-outbreak level | Listeriosis (South Africa 2017-2018) (25) |
| Weekly number of reported cases below the 'epidemic and alert threshold' for eight weeks | Meningitis (Nigeria 2016-2017) (26) |

The use of this criterion to declare the EO, especially in the context of EVD, has been questioned. Firstly, this criterion ignores the possible recrudescence of EVD cases through uncommon transmission routes and persistence of the virus in survivors (sexual transmission, immunocompromised women, and migration) (13–17). Recrudescence of EVD cases has been problematic in the field, as for example, there were three EO declarations in Liberia before the outbreak was actually over (16). Secondly, the use of the maximum incubation period is challenging with typically limited sample sizes available for estimation, and it does not produce any probabilistic risk assessment (18). Lastly, it is important to consider missing cases due to imperfect surveillance during the outbreak and asymptomatic cases that potentially act as invisible transmission sources during the outbreak and could prolong the time to declare the EO (18,19).

Quantitative frameworks which account for these issues are therefore needed to objectively calculate the probability of cases arising in the future to be confident (for example 95% certain) that there will be no further cases after the declaration is made. However, to date, only a small number of studies have focused on developing quantitative methods to define EO criteria, mostly for directly or air-borne transmitted diseases. Nishiura et al. (19) developed a probabilistic method to calculate the probability of observing additional cases of Middle East Respiratory Syndrome (MERS) in the future, based on the distribution of the serial interval (the time between symptom onset in a case and their infector) and the basic reproduction number (the average number of secondary infections generated by a single case in a large population with no immunity) of MERS. Eichner and Dietz (20) used stochastic simulations to determine the length of the case-free period before declaring the extinction of poliovirus with a specified error probability. Thompson et al. (21) used stochastic SEIR (Susceptible-Exposed-Infectious-Recovered) model simulations to assess the influence of underreporting of EVD cases on the confidence of declaring the end of an EVD outbreak.

However, these approaches only address some of the aforementioned issues and have other limitations which we discuss later. Hence, further development of quantitative techniques to define the EO is urgently needed. In this study, we developed a simulation-based method to calculate the probability of cases arising in the future after the outcome of the last detected case. We account for factors that influence the estimated probability such as the underlying reproduction number, the reporting rate and the delay between the onset and the outcome of the last detected case, which for simplicity we will refer to as ‘the onset-to-outcome delay period’. We tested our method on several EVD outbreak scenarios. Finally, we used the simulation results to propose a new quantitative criterion to define the EO for EVD.

## Methods

We developed a quantitative framework to determine the timing of an EO declaration, which divides the outbreak into three phases: the outbreak phase; the onset-to-outcome delay phase; and the EO declaration phase (**Figure 1**). The outbreak phase encompasses the outbreak trajectory which includes all detected cases up until the time that the EO analysis is about to be conducted. The onset-to-outcome delay phase is the period between the onset of the last detected case to the outcome (recovery or death) of that case. In this phase, there is a risk of undetected cases sustaining transmission beyond the last detected case due to underreporting. Accounting for these potential ‘invisible’ sources of transmission is important to determine the EO with confidence, irrespective of potential underreporting. The EO declaration phase starts the day after the outcome of the last case. In this phase, the probability of cases arising in the future is calculated for each day until a predefined probability threshold, here fixed at 5%, is reached.

**Figure 1.**
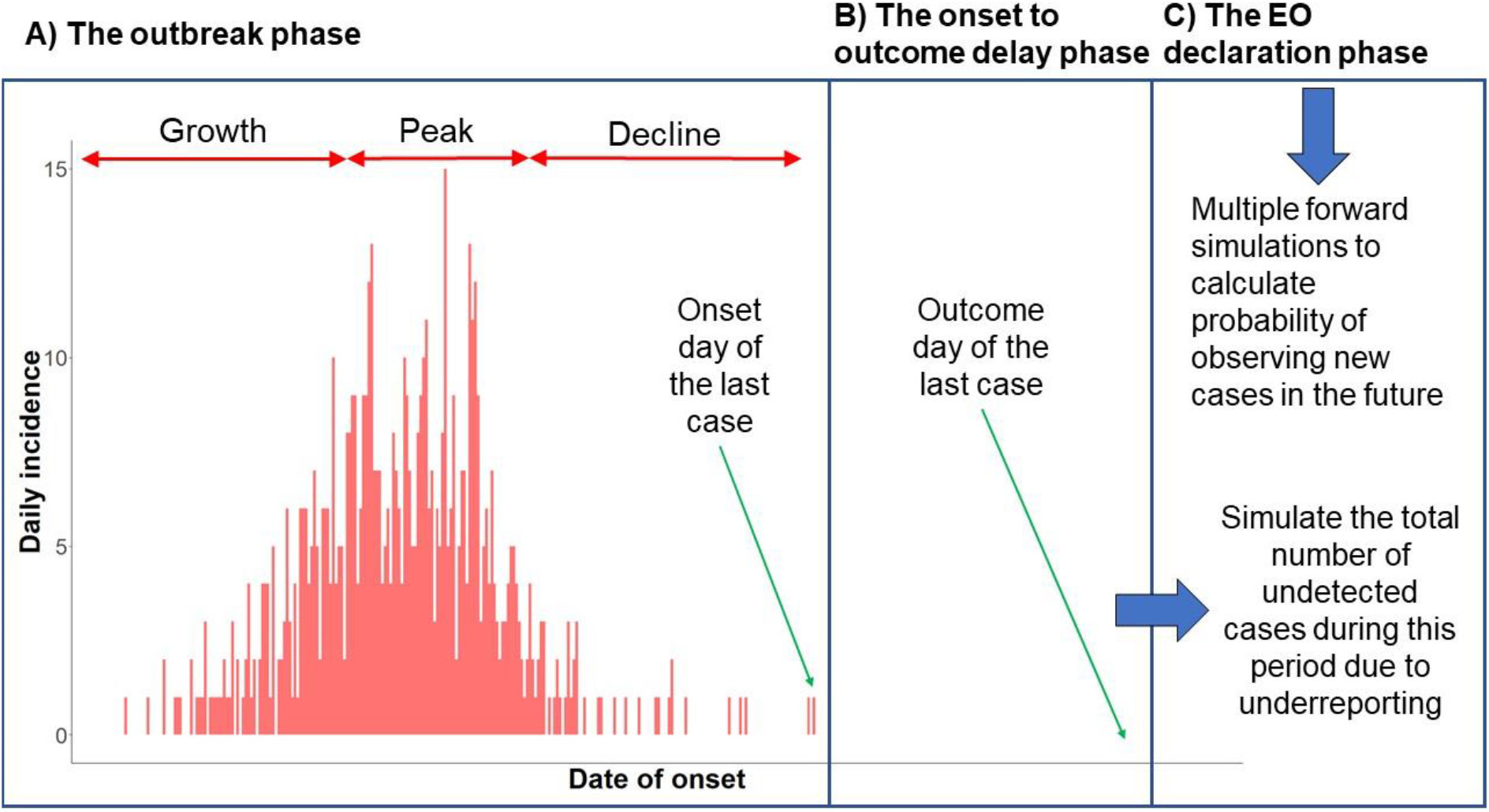
Schematic illustration of the quantitative framework to define the EO. The three main phases of the framework are: A) the outbreak phase (*t* = 1, 2, ⋯, T; T = the observed duration of the outbreak so far); B) the onset-to-outcome delay phase (*d* = 1, 2, ⋯, *D*; *D* = the length of the onset-to-outcome delay period (i.e. recovery or death) of the last detected case); and C) the EO declaration phase (*z* = 1, 2, ⋯, *S*; *S* = the number of days to reach the desired probability threshold of cases arising in the future)

### The outbreak phase

We simulated outbreak data using epidemic parameters from the 2013-2016 EVD epidemic in West Africa (27). Outbreaks were simulated using the *project* function from the *projections* package in the R software, assuming either Poisson or negative binomial offspring distributions to allow for overdispersion (or superspreading, whereby a few cases are responsible for a large proportion of infections) (28). The statistical formulation of the outbreak simulations is as follows: *I*_*t*_ *∼ Poisson*(*R*_*t*_*λ*_*t*_) or *I*_*t*_ *∼ Negbin*(*R*_*t*_*λ*_*t*_, *K*), where *I*_*t*_ is the number of new cases arising (based on symptom onset) at time *t, R*_*t*_ is the instantaneous reproduction number at time *t* (i.e. the average number of secondary infections generated by a case arising at time t, if conditions remain the same), *K* is the overdispersion parameter for the negative binomial distribution and *λ*_*t*_ is the total infectiousness in the population at time *t* which is described by ω and *I*_*t*_ as

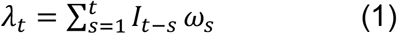

where ω_*s*_ is the typical infectivity profile of a case at time *s*, which can be described by the probability distribution of the serial interval.

The incidence for the first 30 days of the simulated outbreak was taken from the *ebola_sim* line list data from the *outbreaks* R package (29) which is a simulated EVD outbreak data with key properties that match those of the 2013-2016 EVD epidemic in West Africa. After that period, stochastic simulations were run using the model described above. The serial interval was assumed to be gamma distributed with a mean of 15.3 days and a standard deviation of 9.3 days (27). For the negative binomial simulations, the overdispersion parameter *K* was set to values in the range 0.03-0.52, consistent with estimates from the West African EVD epidemic (30,31). We assumed *R*_*t*_ varied over time, with the outbreak divided into three phases: growth, peak, and decline. The ‘growth’ period included the initial 30 days, with an *R*_*t*_ of 1.7 (within the range of Van Kerkhove et al. (27) estimates) and was assumed to last for a total of 90 days. In the ‘peak’ period, assumed to last for 40 days, we used *R*_*t*_ = 1.0. The ‘decline’ period, with *R*_*t*_ values in the range of 0.3-0.9, lasted until the simulated outbreak trajectory had no more cases. For each scenario considered (see simulation scenarios section), 100 stochastic outbreak trajectories were simulated.

### The onset-to-outcome delay phase

It is important to account for the possible new infections caused by undetected cases during the period between the onset and the outcome of the last detected case. We simulated the number of undetected cases in that period using a probabilistic method, under several underreporting assumptions. Using Bayes theorem, an inverse binomial problem was solved to calculate the probability distribution of the total number of cases arising during the onset-to-outcome delay period, given the reporting rate and zero cases detected during that period. The probability mass function of the binomial distribution is described as

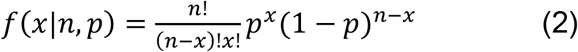

where *n* is the total number of cases (detected and undetected) during the onset-to-outcome delay period, *p* is the reporting rate, and *x* is the number of cases detected during that period, which is zero by definition. Bayes theorem was used to solve the inverse binomial problem of inferring *n*, given the value of *x* and *p*: *f*(*n*|*x, p*) α *f*(*x*|*n, p*)*f*(*n*), with *f*(*n*|*x, p*) the posterior distribution of *n* given the value of *x* and *p, f*(*x*|*n, p*) the binomial likelihood, and *f*(*n*) the prior distribution for *n*.

The prior distribution *f*(*n*) was generated by simulating outbreaks in various hypothetical scenarios with different instantaneous reproduction numbers and reporting rates. For each simulation, several forward trajectories of 21 days starting a day after the onset day of the last case of simulated datasets were generated. Conservatively, a 21-day period was assumed as the maximum onset-to-outcome delay to account for both the average onset-to-death delay and onset-to-discharge delay (upper bound of the onset-to-outcome delay), estimated during the West African EVD epidemic as 8.2 days and 15.1 days respectively with the majority of delays being <21 days (32).

Conditional on the total number of undetected cases during the onset-to-outcome delay period, obtained by solving the inverse binomial problem described above, those cases were allocated probabilistically to each day within this period using a multinomial distribution

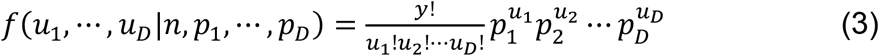

where *y* is the total number of undetected cases obtained by the inverse binomial problem, *u*_1_, ⋯, *u*_*D*_ are the number of undetected cases on day *d* (*d* = 1, 2, ⋯, *D*) after the onset of the last detected case, and *p*_1_, ⋯, *p*_*D*_ are the probabilities of the undetected cases appearing on day *d* (*d* = 1, 2, ⋯, *D*). *p*_1_, ⋯, *p*_*D*_ were calculated by dividing the total infectiousness on each day by the total infectiousness during the whole *D* days period:

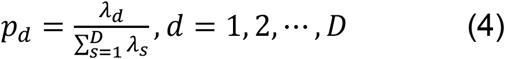

where *λ* is defined as in equation (1). For each simulated outbreak dataset, we simulated the number of undetected cases 10 times. Random allocations were also simulated 10 times for each simulated number of undetected cases.

### The EO declaration phase

In this phase, we performed forward projections of daily incidence of cases, irrespective of reporting status (*Y*_*z*_), for 300 days (*z* = 1, 2, ⋯, 300) following the outcome day of the last detected case based on the combination of the outbreak trajectory and simulated undetected cases during the onset-to-outcome delay phase. Two transmission scenarios were considered: 1) an optimistic transmission scenario where only the last detected case, as well as subsequent undetected cases, can contribute to onwards transmission (illustrative of what would happen if all previous cases have been effectively isolated); and 2) a pessimistic transmission scenario where all cases (detected and undetected) can contribute to onwards transmission, with infectiousness based on the assumed serial interval distribution. To calculate the probability of cases arising in the future on day *z* after the outcome of the last case (*P*_*z*_), for each simulated outbreak dataset, we did 10 forward projections for each combination of simulated number of undetected cases and daily allocations of undetected cases (**Web Figure 1**).

For each projected trajectory, we calculated the total number of projected cases (irrespective of reporting status) from day *z* until 300 days after the outcome of the last case (*C*_*z*_) as

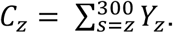

The presence or absence of new cases on day *z* after the outcome of the last case was then summarized as

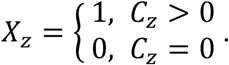

The probability of cases arising in the future on day *z* after the outcome of the last case (*P*_*z*_) was then calculated as

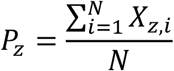

where i = 1, …. N are the different trajectories simulated for a given scenario. We defined the confidence of declaring the EO on day *z* after the outcome of the last case as 1 *− P*_*z*_.

### Simulation scenarios

We simulated several outbreak scenarios with different offspring distributions assuming either optimistic or pessimistic transmission. The offspring distribution parameters in the ‘decline’ period of the outbreak used for the simulations were: 1) Poisson based, *R*_*t*_ = 0.6; 2) Poisson based, *R*_*t*_ = 0.3; 3) Poisson based, *R*_*t*_ = 0.9; 4) Negative binomial based, *R*_*t*_ = 0.6 and *K* = 0.52 (low overdispersion) (31); 5) Negative binomial based, *R*_*t*_ = 0.6 and *K* = 0.18 (medium overdispersion) (30); and 6) Negative binomial based, *R*_*t*_ = 0.6 and *K* = 0.03 (high overdispersion) (31). For the main analyses to calculate *P*_*z*_ and define *Z* where *P*_*z*_ < 5%, the framework used the same *R*_*t*_ value as the simulated outbreak data.

Robustness of the framework to misspecification of *R*_*t*_ value in the ‘decline’ period was tested by sensitivity analyses of framework simulations using different values of *R*_*t*_: 0.3 (underestimation) and 0.9 (overestimation) for simulated outbreak data with *R*_*t*_ = 0.6 in the ‘decline’ period. **Web Table 1** shows the complete combination of all simulation scenarios explored.

## Results

Using the quantitative simulation framework developed, we calculated the probability of cases arising in the future on various numbers of days after the outcome of the last detected case. We considered different scenarios (summarized in **Web Table 1**) by varying the offspring distribution of the outbreak, the length of the onset-to-outcome delay period, reporting rate, and assumed transmission scenario. **Figure 2** shows the number of days taken (from the outcome of the last detected case) for the probability of cases arising in the future to fall below 5%, for six different offspring distributions.

**Figure 2.**
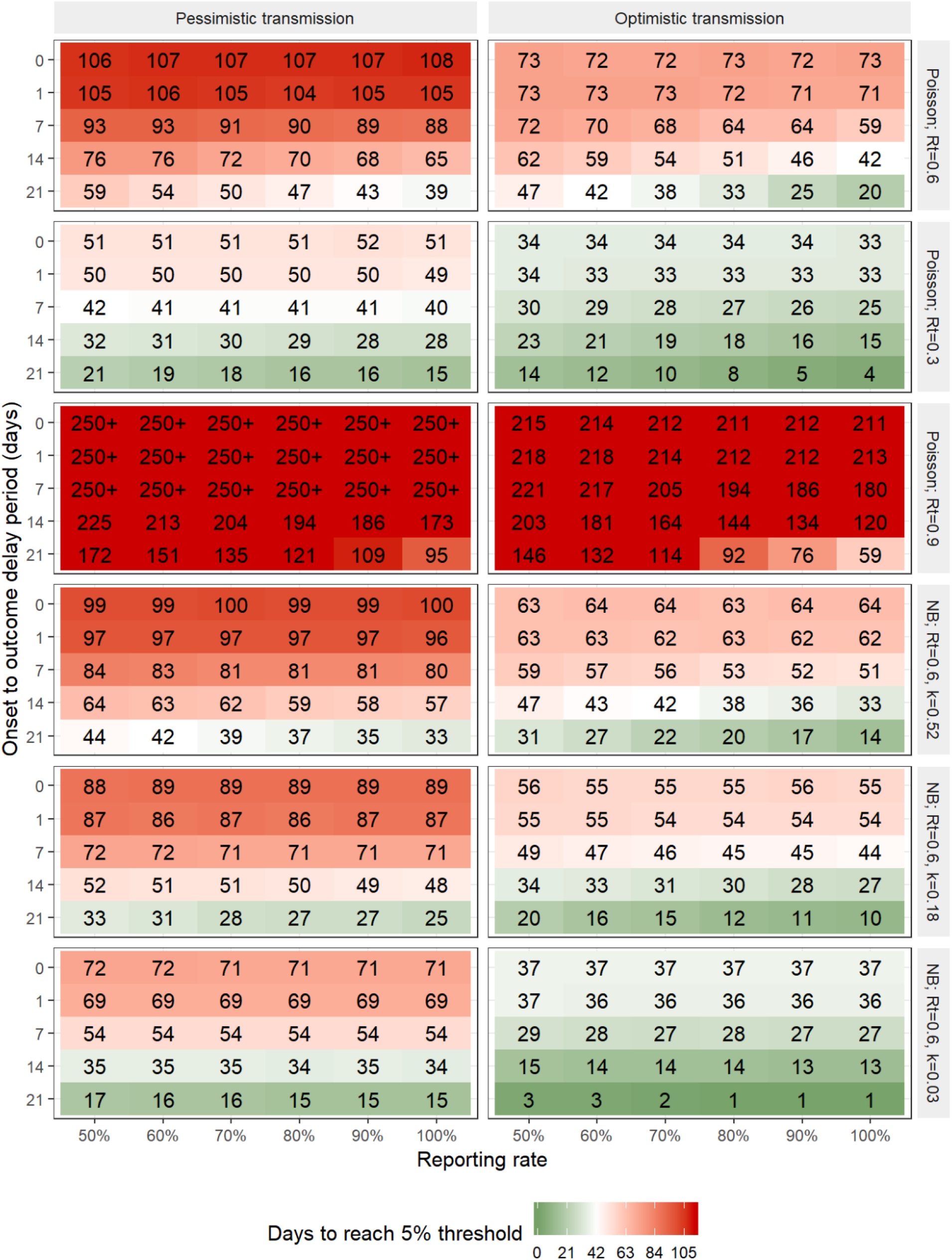
Number of days (from the outcome of the last detected case) until the probability of cases arising in the future falls below the threshold 5% for various offspring distributions during the ‘decline’ period (from the top row to the bottom row: 1) Poisson based, *R*_*t*_ = 0.6; 2) Poisson based, *R*_*t*_ = 0.3; 3) Poisson based, *R*_*t*_ = 0.9; 4) Negative binomial based, *R*_*t*_ = 0.6 and *K* = 0.52; 5) Negative binomial based, *R*_*t*_ = 0.6 and *K* = 0.18; and 6) Negative binomial based, *R*_*t*_ = 0.6 and *K* = 0.03) and for the pessimistic (left) and optimistic (right) transmission scenarios, as a function of the length of the reporting rate and the onset-to-outcome delay period. An onset-to- outcome delay of zero days corresponds to counting days from the onset day of the last detected case. Red cells denote longer waiting times to reach the 5% probability threshold while green cells denote shorter waiting times. Current WHO criterion is 42 days after the outcome of the last detected case.

Our simulations show that the underlying offspring distribution during the ‘decline’ period of the outbreak is the most determinant factor in how long it takes for the probability of cases arising in the future dropped below 5%. The waiting time is longer when the value of the *R*_*t*_ is higher. On the other hand, a higher level of overdispersion in the offspring distribution leads to shorter waiting times to reach <5% probability of cases arising in the future. Outbreaks with no overdispersion tend to have a consistent outbreak length and total number of cases during the outbreak. However, outbreaks with high overdispersion are often shorter with a smaller final epidemic size, but by chance, can also be long with a very large number of cases (**Web Figure 2**).

The length of the onset-to-outcome delay period is also shown to affect the waiting time to reach <5% probability of cases arising in the future; with longer lengths of the onset-to-outcome delay period leading to shorter waiting times. On the other hand, the influence of the reporting rate is only major when there is a long onset-to-outcome delay period; with higher reporting rates leading to shorter waiting times. When the onset day was used as the reference of the waiting time to reach <5% probability of cases arising in the future (length = 0), the waiting times are constant. For example, the waiting time for Poisson distribution-based outbreaks with *R*_*t*_ = 0.6 for all tested values of reporting rate are around 107 and 72 days for pessimistic and optimistic scenario respectively (**Figure 2**).

The assumption on whether past cases can still contribute to transmission or not (pessimistic or optimistic transmission) also substantially affects the waiting time; as expected, assuming perfect isolation of past cases leads to shorter waiting times. Finally, sensitivity analyses suggested that the developed framework was not robust to misspecification of the value of the reproduction number in the ‘decline’ period (**Web Figure 3 & 4**).

**Figure 3.**
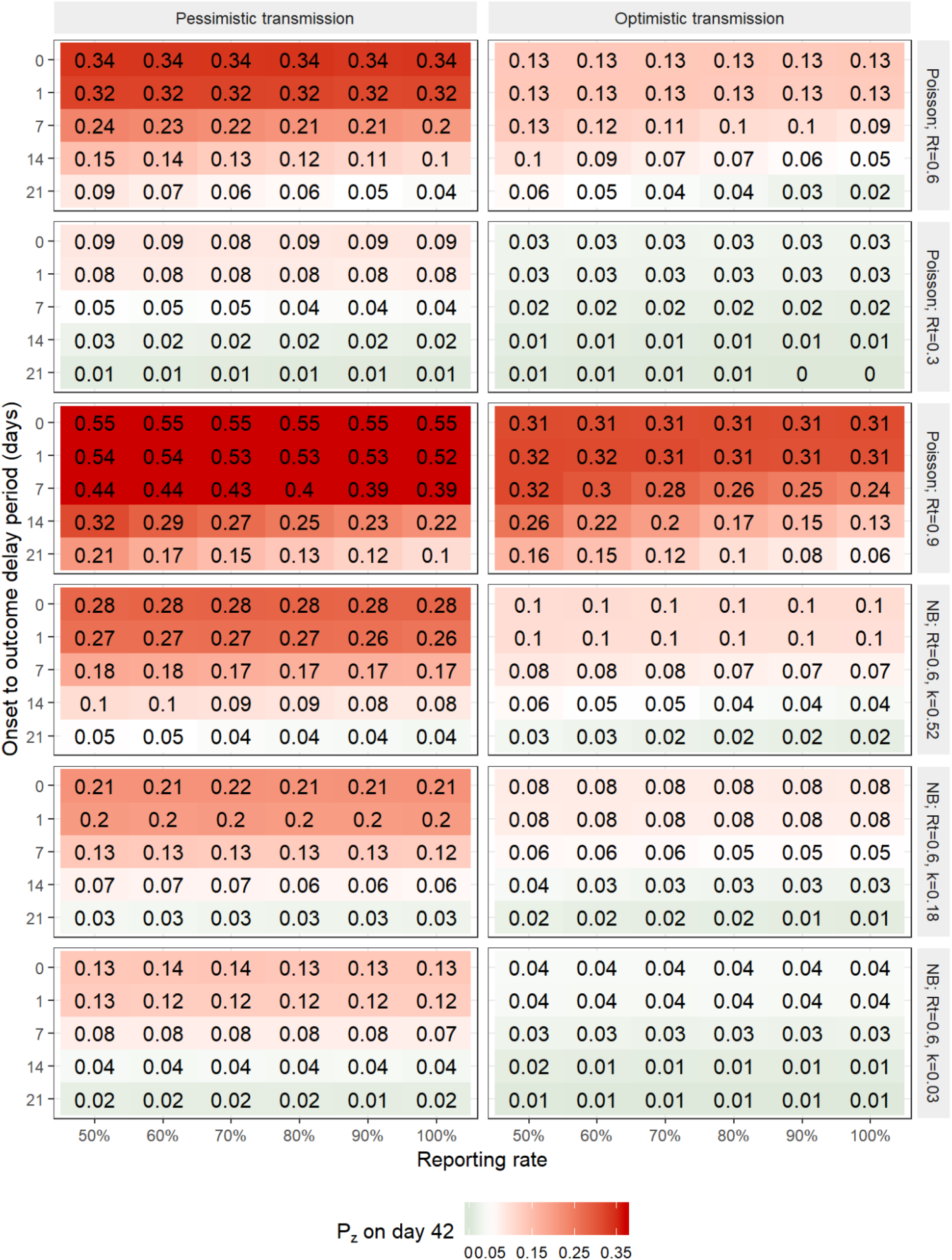
Probability of cases arising more than 42 days after the outcome of the last detected case for various offspring distributions during the ‘decline’ period (from the top row to the bottom row: 1) Poisson based, *R*_*t*_ = 0.6; 2) Poisson based, *R*_*t*_ = 0.3; 3) Poisson based, *R*_*t*_ = 0.9; 4) Negative binomial based, *R*_*t*_ = 0.6 and *K* = 0.52; 5) Negative binomial based, *R*_*t*_ = 0.6 and *K* = 0.18; and 6) Negative binomial based, *R*_*t*_ = 0.6 and *K* = 0.03) and for the pessimistic (left) and optimistic (right) transmission scenarios, as a function of the length of the reporting rate and the onset-to-outcome delay period. An onset-to-outcome delay of zero days corresponds to counting days from the onset day of the last detected case. Red cells denote higher probability of cases arising more than 42 days after the outcome of the last detected case while green cells denote lower probability.

We calculated the probability of cases arising after 42 days, following the outcome of the last detected case (**Figure 3**), which is the current WHO criterion to declare the EO for EVD. Our simulations show that in most scenarios considered, the current WHO criterion corresponds to a probability of cases arising in the future well above 5%. The corresponding probability is <5% only when the onset-to-outcome delay period is very long (at least two or three weeks, which is unusual) and the reporting rate is high, although with low *R*_*t*_ value, high overdispersion or optimistic transmission the probability reaches <5% in more parameter combinations. Once again, misspecification of the value of *R*_*t*_ in the ‘decline’ period strongly affected these results (**Web Figure 5 & 6**). An additional analysis, accounting for the 90-day period of enhanced surveillance after the EO declaration of EVD (8) shows that the probability of cases arising in the future for most of simulation scenarios are <5% at the end of the 132-day period (42 + 90 days) except when the value of *R*_*t*_ is high (**Web Figure 7**).

## Discussion

We have developed a simulation-based framework to calculate the probability of cases arising in the future after the outcome day of the last detected case of an outbreak. Our simulations show that the reporting rate and the delay between the onset and the outcome of the last detected case are important factors that need to be considered in assessing the EO. We applied this simulation-based framework to analyze a range of simulated EVD outbreaks with different levels of superspreading consistent with EVD epidemiology, and different scenarios spanning extreme assumptions about the effectiveness of case isolation (no case isolation or pessimistic transmission to perfect case isolation or optimistic transmission).

Our results show that under the current WHO criterion to declare the EO for EVD of 42 days with no cases after the last detected case dies or tests negative for the virus twice, the probability of cases arising in the future is still high (up to 54%) in most scenarios. Thus, a more robust EO criterion, supported by quantitative evidence, is needed to minimize the risk of flare-ups of cases after EO declaration. This was also highlighted by the multiple flareups of cases after EO declarations were made at the tail end of the West African EVD epidemic (33).

Some of the flare-ups happened 51, 68, 78, and 80 days after the EO declarations, highlighting the importance of the WHO recommendation of 90 days of enhanced surveillance after the 42-day period of waiting time from the outcome of the last detected case to declare the EO of EVD. The importance of this enhanced surveillance period is also supported by our results. Our simulations show that the probability of cases arising in the future after this 132-day (42 + 90 day) period from the outcome of the last detected case is generally lower than 5% except in rare scenarios with a high reproduction number during the ‘decline’ phase (**Web Figure 2**).

Our simulations show that the estimated probability of cases arising in the future were very sensitive to the value of *R*_*t*_, the instantaneous reproduction number at time *t*, during the ‘decline’ period. Therefore, monitoring *R*_*t*_ during an outbreak is very important to accurately define the EO. Especially in the ‘decline’ period, ensuring that *R*_*t*_ is reduced well below 1 is critical for bringing the outbreak to an end. Current methods available to estimate the instantaneous reproduction number during the outbreak may suffer from imprecision if there is uncertainty in the serial interval distribution estimates and more generally at the EO when case numbers are low (34). Our simulations show that the framework we developed is sensitive to the instantaneous reproduction number estimate. These factors combined emphasize the need for continued assessment of the instantaneous reproduction number throughout the outbreak and particularly as case numbers decrease. Consequently, it is hard to define a single criterion for the EO given how influential *R*_*t*_ is.

We found that the duration of the onset-to-outcome delay period had a large impact on the waiting time to declare the EO, with a short onset-to-outcome delay leading to a longer time to declare the EO. Given the potential variability in this delay period between cases, in particular depending on outcome (mean onset-to-recovery and onset-to-death were estimated as 14.4-15.3 and 6.2-8.8 days respectively (32)), our results do not support the current WHO single criterion to declare the EO, irrespective of the last detected case’s outcome.

Our study also shows that the reporting rate plays an important role in assessing the EO, and that, in line with results by Thompson et al. (21), a low reporting rate would lead to a lower confidence in declaring the end of an EVD outbreak at day 42 after the outcome of the last case, i.e. using the current WHO criterion. Hence, for outbreaks with low reporting rates, a longer waiting time would be needed to declare the EO. However, we found that the dependency on the reporting rate became negligible as the onset-to-outcome delay period decreases. This suggests that using the onset day, rather than the outcome day, as the baseline of the waiting time should be considered as it may be more robust to variability in the outbreak context. This would also account for the three possible scenarios for the last detected case which could include any delays in testing of the last detected case or delays in burial (8).

We therefore recommend a shift to a preliminary end-of-outbreak declaration after 63 days from the symptom onset day of the last detected case. This should be followed by a 90-day enhanced surveillance, after which the official end-of-outbreak can be declared. This corresponds to less than 5% probability of flare ups in most of the scenarios examined in our study.

Our simulation-based framework was tested under multiple assumptions regarding superspreading or overdispersion. Our simulations show that the waiting time decreases as overdispersion increases. Exploring the optimistic and pessimistic transmission scenarios, we found that the current WHO criterion will perform best in the optimistic transmission scenario. However, given the difficulty of controlling and isolating cases during an outbreak, this optimistic scenario should be considered a best-case scenario and the pessimistic scenario a worst-case scenario.

We propose a quantitative framework that can be used to assess the EO. The first step is estimating the key outbreak parameters: instantaneous reproduction number, reporting rate, and serial interval distribution. These can be estimated from outbreak data using widely available and established methods which increasingly account for sparse data (34–37). The next step is to implement the method developed in this study to determine the day when the probability of cases arising in the future is deemed acceptable (in this study < 5%), and an outbreak can be declared over. The developed method is generic; thus it could be implemented for outbreaks of other pathogens, primarily if they are air-borne or transmitted by direct contact.

Based on our simulation results, we recommend a new criterion using the day of onset of the last detected case as the baseline of the waiting time rather than the outcome day. The onset day is usually captured better and is less affected by diagnostic waiting times. Thus, it is also generalisable to other diseases. However, given the sensitivity of the probability of cases arising in the future to the value of *R*_*t*_, it is difficult to suggest a single criterion for EO irrespective of *R*_*t*_. Based on our results we therefore make three general recommendations: i) a shift from counting down to EO from 42 days from the day of outcome to 63 days from the day of onset of the last detected case leading to a preliminary EO declaration; ii) emphasis on the importance of properly resourcing the enhanced 90-day surveillance post-EO declaration to ensure that any flareups are quickly detected and controlled before the final EO declaration (63 + 90 days after the onset of the last detected case); and iii) regular estimation and re-estimation of the reproduction number particularly in the decline phase.

Finally, some aspects of EVD transmission were not covered by our simulation framework, which deserve to be highlighted. The framework developed did not consider additional cases that arise from less common transmission routes such as migration, sexual transmission, and immunocompromised pregnant women. These caveats make the current policy to keep active case detection up to 90 days after the EO declaration essential if the framework and model were to be adopted. Although the framework accounts for superspreading (by allowing overdispersion in the offspring distribution), we did not explicitly model superspreading events in the context of unsafe burial practices that lead to large-scale funeral exposures (30,38). Nevertheless, the work presented here demonstrates the value of the development of a quantitative framework than can be used to help decision-makers to objectively assess the risk of flare-ups of cases after the EO declaration. It also highlights the limitations of the current WHO criterion for declaring the EO of EVD.

## Data Availability

The simulated datasets used in the study are available on the github repository provided. No primary data were used.

https://github.com/andradjaafara/defining_the_end_of_an_outbreak

## Acknowledgements

Author affiliation: MRC Centre for Global Infectious Disease Analysis, Imperial College London, London, United Kingdom (Bimandra Adiputra Djaafara, Natsuko Imai, Christl Ann Donnelly, Anne Cori); Ejikman-Oxford Clinical Research Unit, Jakarta, Indonesia (Bimandra Adiputra Djaafara); World Health Organization, Regional Office for Africa, Brazzaville, Congo (Esther Hamblion, Benido Impouma); Department of Statistics, University of Oxford, Oxford, United Kingdom (Christl Ann Donnelly).

BAD acknowledges a matched MRC Centre 1+3 studentship funding by Imperial College London School of Public Health. NI, CAD and AC acknowledge joint centre funding from the UK Medical Research Council and Department for International Development.

Conflict of interest: None declared.

## Abbreviations

DRC: Democratic Republic of the Congo
EO: end of an outbreak
EVD: Ebola Virus Disease
MERS: Middle East respiratory syndrome
SEIR: Susceptible – Exposed – Infectious – Recovered
WHO: World Health Organization

